# Normobaric Hyperoxia Combined with Endovascular Treatment Based on Temporal Gradient: A dose-escalation study

**DOI:** 10.1101/2023.12.05.23299563

**Authors:** Weili Li, Sifei Wang, Lan Liu, Jiahao Chen, Jing Lan, Jiayue Ding, Zhiying Chen, Shuhua Yuan, Zhifeng Qi, Ming Wei, Xunming Ji

**Author notes:** Weili Li and Sifei Wang contributed equally to this article. **Corresponding author:** Dr. Xunming Ji, Department of Neurosurgery, Xuanwu Hospital, Capital Medical University, Beijing, China.; Dr. Ming Wei, Department of Neurosurgery, Tianjin Huanhu Hospital, Tianjin, China.

## Abstract

**BACKGROUND:** Normobaric hyperoxia (NBO) has neuroprotective effects in acute ischemic stroke (AIS). Thus, we aimed to identify the optimal NBO treatment duration combined with endovascular treatment (EVT).

**METHODS:** Patients with acute stroke who had an indication for EVT at Tianjin Huanhu Hospital were included and randomly assigned to four groups (1:1 ratio) based on NBO therapy duration: 1) Sham-NBO group (oxygen 1 L/min continuously for 4 h); 2) NBO-2h group (10 L/min continuously for 2 h); 3) NBO-4h group (10 L/min continuously for 4 h); and 4) NBO-6h group (10 L/min continuously for 6 h). The primary outcome was cerebral infarction volume at 72 h after randomization. The primary safety outcome was the 90-d mortality rate.

**RESULTS:** A total of 100 patients were included (Sham-NBO group, n=25; NBO-2h group, n=25; NBO-4h group, n=25; and NBO-6h group, n=25). The 72-h cerebral infarct volumes were 39.4 ± 34.3 ml, 30.6 ± 30.1ml, 19.7 ± 15.4 ml, and 22.6 ± 22.4 ml, respectively (P=0.013). The NBO-4h and NBO-6h groups exhibited significant differences compared to the Sham-NBO group (adjusted P values: 0.011 and 0.027, respectively). No significant differences were found between the NBO-4h and NBO-6h groups. The National Institute of Health Stroke Scale (NIHSS) scores at 24 h, 72 h, and 7 d, and the changes in NIHSS scores from baseline to 24 h were significantly different in the NBO-4h and NBO-6h groups compared with the Sham-NBO group (P<0.05). No significant differences were observed between the NBO-4h and NBO-6h groups in the NIHSS assessments. No significant differences were noted among groups in the 90-d mortality rate, symptomatic intracranial haemorrhage, early neurological deterioration, and severe adverse events.

**CONCLUSIONS:** The effectiveness of NBO therapy was associated with oxygen administration duration. In patients with AIS who undergone EVT, NBO treatment for 4-6 h may yield better outcomes than other oxygen therapy regimens or low flow oxygen therapy.

**REGISTRATION:** URL: https://www.clinicaltrials.gov; Unique identifier: NCT05404373.

## INTRODUCTION

The estimated prevalence of stroke among Chinese adults is 39.3%, which is the highest stroke prevalence worldwide.^1^ This prevalence is further exacerbated by the ongoing aging of the population, posing a substantial economic burden on both China’s economy and social development. Reducing stroke incidence and enhancing post-stroke disability management are crucial. Notably, among stroke patients, a significant 88% are afflicted by acute ischemic stroke (AIS), primarily resulting from acute vascular occlusion.^2^

Currently, the most effective treatment for AIS is to recanalize the occluded blood vessels.^3^ With technological advancements, the success rate of vascular recanalization has surpassed 80%. However, despite these improvements, the rate of favourable outcomes for patients remains below 50%.^4,5^ This disparity can be attributed to several factors, including the limited salvageable ischemic penumbra during vascular recanalization and the occurrence of reperfusion injury following the reopening of the vessel.^6–8^

Considering these challenges, we proposed a multi-stage, sequential treatment approach that builds upon vascular recanalization. This approach aims to optimize protection of the ischemic penumbra before recanalization and to mitigate reperfusion injury after the vessel is reopened.^9^

Presently, the optimal therapeutic approach for AIS involves a combination of vascular recanalization and neuroprotective treatment.^10–12^ Normobaric hyperoxia (NBO) is considered a promising neuroprotective therapy due to its rapid onset, minimal adverse effects, ease of administration, cost-effectiveness, and wide applicability. Basic and clinical research conducted by our team has demonstrated significant neuroprotective benefits of normobaric hyperoxia.^13–15^ Specifically, its ability to enhance oxygen levels in ischemic regions, mitigate blood-brain barrier damage, enhance neurological function scores, and reduce cerebral infarct volumes, leading to improved clinical outcomes. However, no consensus is currently available in clinical studies on the optimal duration of NBO treatment. NBO treatment has been administered for 8 h in patients who have not undergone revascularisation therapy,^16^ and in patients before revascularisation.^17^ NBO treatment has also been administered for 6 h after revascularisation.^18^ To the best of our knowledge, the optimal duration of NBO treatment remains unreported.

The primary objective of this study was to investigate the optimal duration of NBO treatment by employing sequential time gradients. This research strategy sought to achieve a balance between minimizing the potential side effects of NBO treatment and maximizing its therapeutic advantages. The aim was to generate empirical evidence that can inform and promote the future clinical application of NBO therapy.

## METHODS

### Study design

This single-centre, randomized controlled dose-escalation study with a blinded endpoint was designed to evaluate the impact of varying durations of NBO therapy on the efficacy and safety of treatment for AIS. And it was conducted from June 2022 and september 2023 at Tianjin Huanhu Hospital in China. The Trials flowchart is shown in Figure 1.

**Figure 1.**
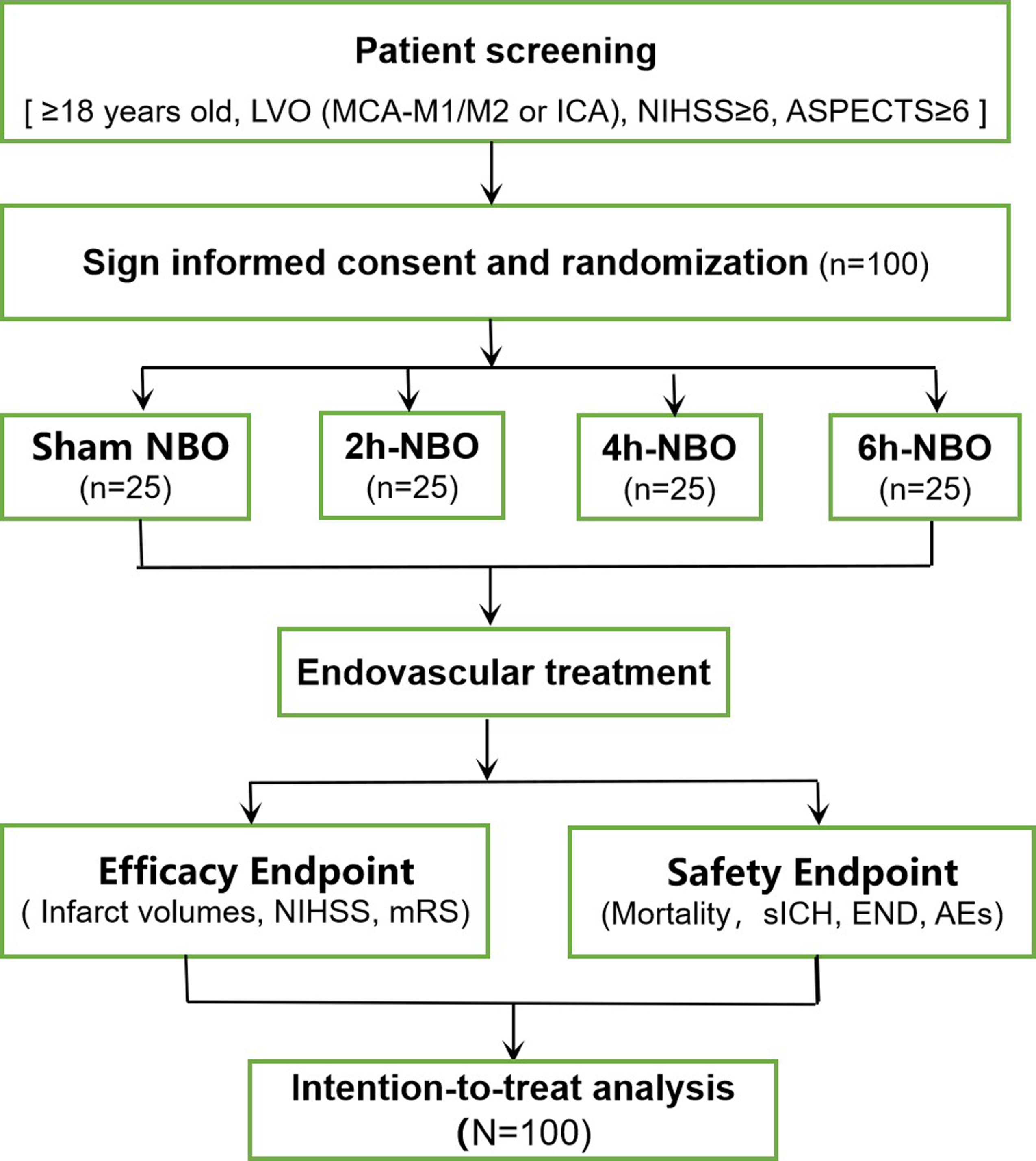
Study design diagram. LVO, large vessel occlusion; MCA, middle cerebral artery; ICA, internal carotid artery; NIHSS, national institutes of health stroke scale; ASPECTS, Alberta Stroke Program Early CT Score; NBO, normobaric hyperoxia; mRS, modified Rankin scale; sICH, Symptomatic Intracranial Hemorrhage; END, Early neurological deterioration; AEs, adverse events.

The study was approved by the Ethics Committee of Huanhu Hospital, Tianjin Medical University (Approval No. 2022-076), and is registered on Clinicaltrials.gov with the Unique Protocol ID: TD-NBO and Identifier: NCT05404373.

An independent steering committee, consisting of independent researchers, physicians, and statisticians, oversaw the design and implementation of the research. The committee members were responsible for ensuring the meticulous execution of the study, data collection, and subsequent statistical analysis. Prior to patient enrolment, the personnel involved in the study underwent rigorous research and procedural training, including simulated training, to ensure the highest standards of research quality. All patients or their legal representatives provided informed consent before participating in the study.

### Patient selection

The inclusion criteria were as follows: patient age ≥ 18 years, symptoms and signs consistent with acute anterior circulation ischemic stroke, a National Institutes of Health Stroke Scale (NIHSS) score of ≥ 6, and an Alberta Stroke Program Early CT Score (ASPECTS) of ≥ 6. The criteria for eligibility for endovascular treatment (EVT) were as follows: time from symptom onset to randomisation within 24 h; consciousness level assessed by the NIHSS with a score of 0 or 1; pre-stroke modified Rankin Scale (mRS) score of 0-1; confirmation of large vessel occlusion through pre-randomization computed tomography angiography or magnetic resonance angiography, showing occlusion in the internal carotid artery or middle cerebral artery M1/M2 segments; informed consent provided by the patient or their legal representatives.

The following exclusion criteria were applied: rapid neurological improvement; an NIHSS score < 10 or evidence of spontaneous vessel recanalization; active bleeding or a known bleeding tendency; presence of seizures during the stroke episode preventing accurate NIHSS scoring; platelet count < 100×10^9 /L; known hereditary or acquired bleeding diathesis, coagulation factor deficiency, or recent use of anticoagulant drugs (international normalized ratio > 3 or partial thromboplastin time > 3 times normal); evidence of cardiac, hepatic, or renal dysfunction; baseline blood glucose level < 50 mg/dL (2.78 mmol) or > 400 mg/dL (22.20 mmol); uncontrolled hypertension (systolic blood pressure > 185 mmHg; diastolic blood pressure > 110 mmHg); expected survival time < 90 days; pregnancy; chronic obstructive pulmonary disease, pulmonary inflammation, pleural effusion, acute respiratory distress syndrome, or irregular respiration; patients with unstable vital signs (heart rate ≤ 50 beats/minute or ≥ 120 beats/minute, oxygen saturation ≤ 90%, respiratory rate ≥ 30 breaths/minute or ≤ 10 breaths/minute); patients unable to complete a 90-day follow-up of any reasons; other urgent medical conditions requiring oxygen therapy; upper gastrointestinal bleeding or the presence of nausea, vomiting, or other conditions that hinder the use of a face mask for oxygen therapy.

### Randomization and blinding method

Patients were allocated randomly in a 1:1 ratio to one of four groups: Sham-NBO group, NBO-2h group, NBO-4h group, and NBO-6h group, utilizing a completely randomized design facilitated by the Interactive Web Response System.The randomization code is generated by a specially trained physician who logs into the randomization system. This physician then informs the study practitioner whether the patient is assigned to the NBO or low-flow oxygen group. This study employed an open-label design, with no blinding during the administration of oxygen treatment. However, a blinded approach was rigorously implemented for the assessment of outcomes. Specifically, blinding measures were implemented for the imaging assessors, clinical outcome assessors, and follow-up personnel. The statistician responsible for data analysis was also blinded. Throughout the experiment, strict protocols were in place to prevent non-blinded personnel from participating in clinical assessments or engaging in communication with the blinded assessors.

### NBO protocol

Upon randomization, initiating oxygen therapy promptly is crucial, with a strict time limit of no later than 30 min post-randomization. NBO treatment was administered through inhalation of 100% oxygen at a flow rate of 10 L/min, utilizing a storage oxygen mask. According to group assignment, patients underwent NBO therapy for either 2, 4, or 6 h. Conversely, patients in the Sham-NBO group were treated with oxygen at a rate of 1 L/min through a mask for a duration of 4 h. To ensure comprehensive monitoring, blood gas analysis was conducted for all patients before the completion of their respective oxygen therapy sessions.

Additionally, local anaesthesia was administered to patients whenever possible. However, if a patient became agitated and uncooperative, switching to general anaesthesia was considered. In such instances, the method of oxygen delivery was modified from mask oxygen delivery to intubation for oxygen administration. In this scenario, for a flow rate of 10 L/min, the method was adjusted to deliver an inspiratory oxygen fraction (FiO2) of 1.0. Similarly, for a flow rate of 1 L/min, the FiO2 was adjusted to 0.3 if intubation was performed. If a patient still required supplemental oxygen to maintain oxygen saturation levels above 94% despite following the prescribed oxygen administration protocol, the use of a nasal cannula for oxygen delivery was considered. In such cases, the oxygen flow rate did not exceed 3 L/min.

### Endovascular Treatment (EVT)

All patients should receive EVT to facilitate recanalization of the occluded vessel. EVT included mechanical thrombectomy, thrombus aspiration, or a combination of both, with stent implantation if necessary. Furthermore, if a patient met the eligibility criteria for intravenous thrombolysis prior to undergoing EVT, alteplase was administered for intravenous thrombolysis at a dosage of 0.9 mg/kg body weight, as a bridging therapy before commencing EVT.

### Study outcomes

The primary outcome was the volume of cerebral infarction 72 h after treatment, evaluated by magnetic resonance imaging - diffusion weighted imaging (MRI-DWI). The core infarct was defined as the region of apparent diffusion coefficient (ADC) values <620×10^−6^ mm^2^/s.^19^ We measured infarct volumes using eStroke software (https://www.brainomix.com/stroke/) ^20^

The following secondary efficacy outcomes were recorded: modified Rankin Scale (mRS) score at 90 d; the proportion of patients achieving mRS scores of 0-2 at 90 d; partial pressure of oxygen (PaO2) following oxygen therapy; the NIHSS scores at various time intervals (24 h, 72 h, and 7 d); changes in the NIHSS score from baseline to 24 h; and successful vessel recanalization, defined as grade 2b, 2c, or 3 on the extended Thrombolysis in Cerebral Infarction scale (eTICI, 0-3, with higher grades indicating increased reperfusion).^21^

The mRS scores were obtained through structured telephone follow-ups conducted by personnel who had undergone specialized training and were unaware of the treatment group assignments. Conversely, NIHSS scores were evaluated on-site by assessors with a certification for NIHSS score assessment.

The following safety outcomes were evaluated: the 90-d mortality rate, 90-d stroke-related mortality rate, incidence of any intracranial haemorrhage, incidence of symptomatic intracranial hemorrhage (sICH), early neurological deterioration (END), incidence of severe adverse events (SAEs), and incidence of pneumonia.

sICH was defined according to the European Cooperative Acute Stroke Study III criteria as any type of intracranial hemorrhage associated with an increase of at least 4 points in the NIHSS score or death and was assessed as being the predominant cause of neurologic deterioration.^22^

END was defined as an increase of ≥4 points in the NIHSS score 24 h after thrombectomy compared with the baseline NIHSS score.^23^ SAEs were characterized as events resulting in a deterioration of the patient’s condition, prolonged length of hospital stay, life-threatening situations, or death, which specifically included early neurological deterioration, symptomatic intracranial haemorrhage, 90-d mortality, re-occlusion of a recanalized blood vessel, increase in the infarct volume, recurrent stroke, severe pulmonary infection, and deep venous thrombosis of lower limbs.

### Statistical Analysis

This study adopted an intention-to-treat analysis approach. Continuous variables were presented as mean (standard deviation, SD) or median (interquartile range, IQR), while categorical data were presented as counts and percentages. Descriptive statistical analysis was used for data on baseline characteristics. To address missing data, multiple imputation techniques were applied.

For continuous variables with a normal distribution, one-way analysis of variance was used. In cases where continuous variables do not adhere to a normal distribution, non-parametric tests were applied. In the event of observed differences between the groups, pairwise comparisons were conducted. To mitigate the risk of type II errors due to multiple comparisons, adjusted p-values were used. For comparisons involving proportions across multiple groups, chi-squared tests and Fisher’s exact tests were employed as appropriate. A P-value <0.05 was considered statistically significant.

The study was analysed in compliance with standard Good Clinical Practice regulations, database, and the statistical analysis plan was locked before unblinding. Analyses were performed using SPSS software for Windows (Version 26.0. Armonk, NY: IBM Corp).

### Sample Size Calculation

The sample size was determined based on the literature and pre-experimental results.^14^ Under the assumption that the dose response on the infarct volume is monotone but not necessarily equal among NBO groups, we used William’s test to calculate the minimum effective dose. We considered 4 groups: control group (sham NBO), NBO for 2 hours, 4 hours, and 6 hours. The expected effect size was 15 ml between the NBO and sham NBO group. With a type I error α= 0.05 and a power 1 −β= 0.8, the required sample size was 25 per group with a total of 100 patients.

## RESULTS

### Baseline characteristics

This study enrolled 100 patients between June 2022 and September 2023 with 25 patients in each group. Baseline characteristics included age, sex, medical history, admission NIHSS score, ASPECT score, baseline oxygen tension levels, infarct volume, location of occluded vessels, Trial of ORG 10172 in Acute Stroke Treatment (TOAST) classification, and time intervals from onset to randomization and from oxygen inhalation to recanalization. No significant differences were observed in patient baseline clinical characteristics among the four groups (P > 0.05, Table 1).

**Table 1.**
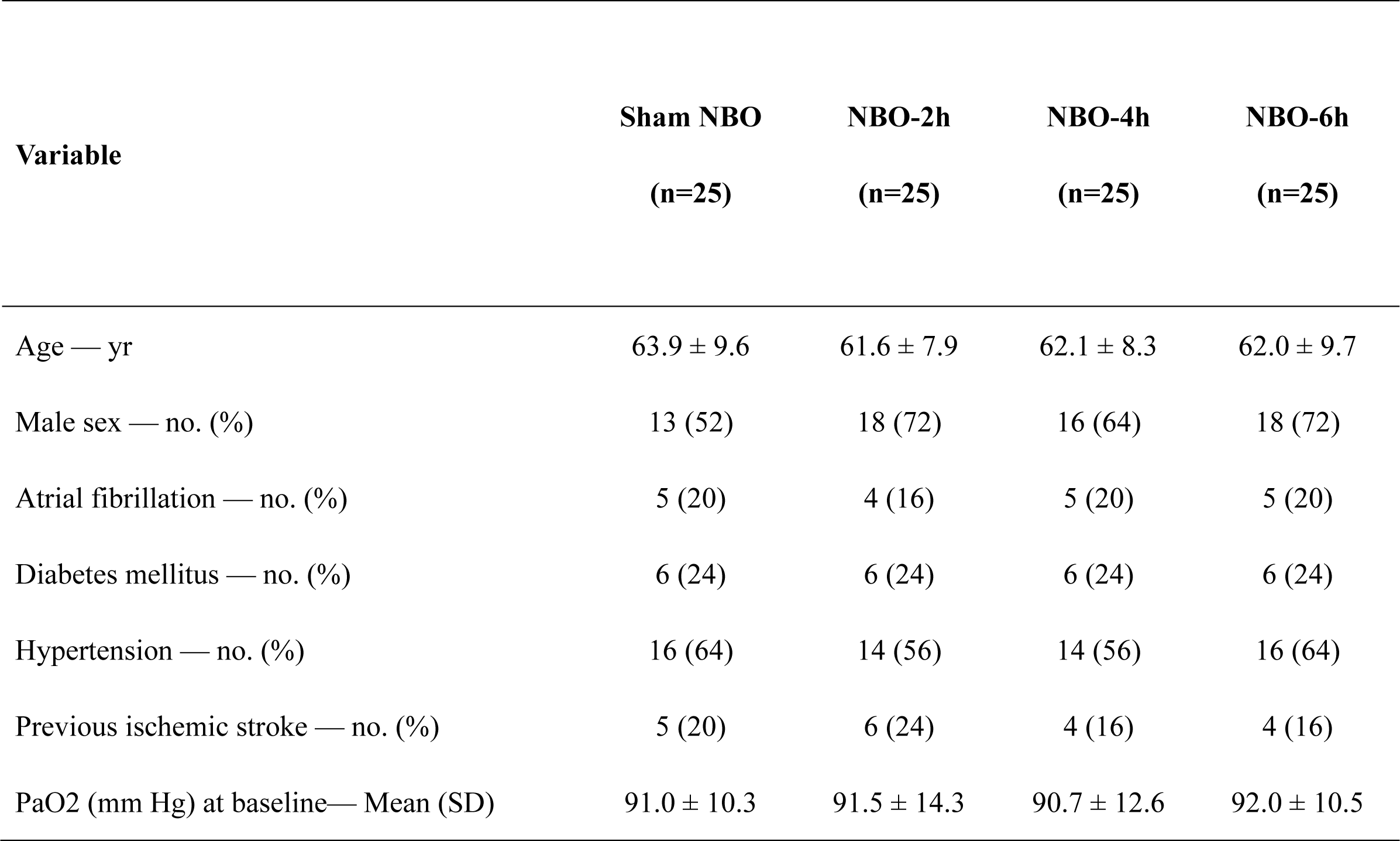
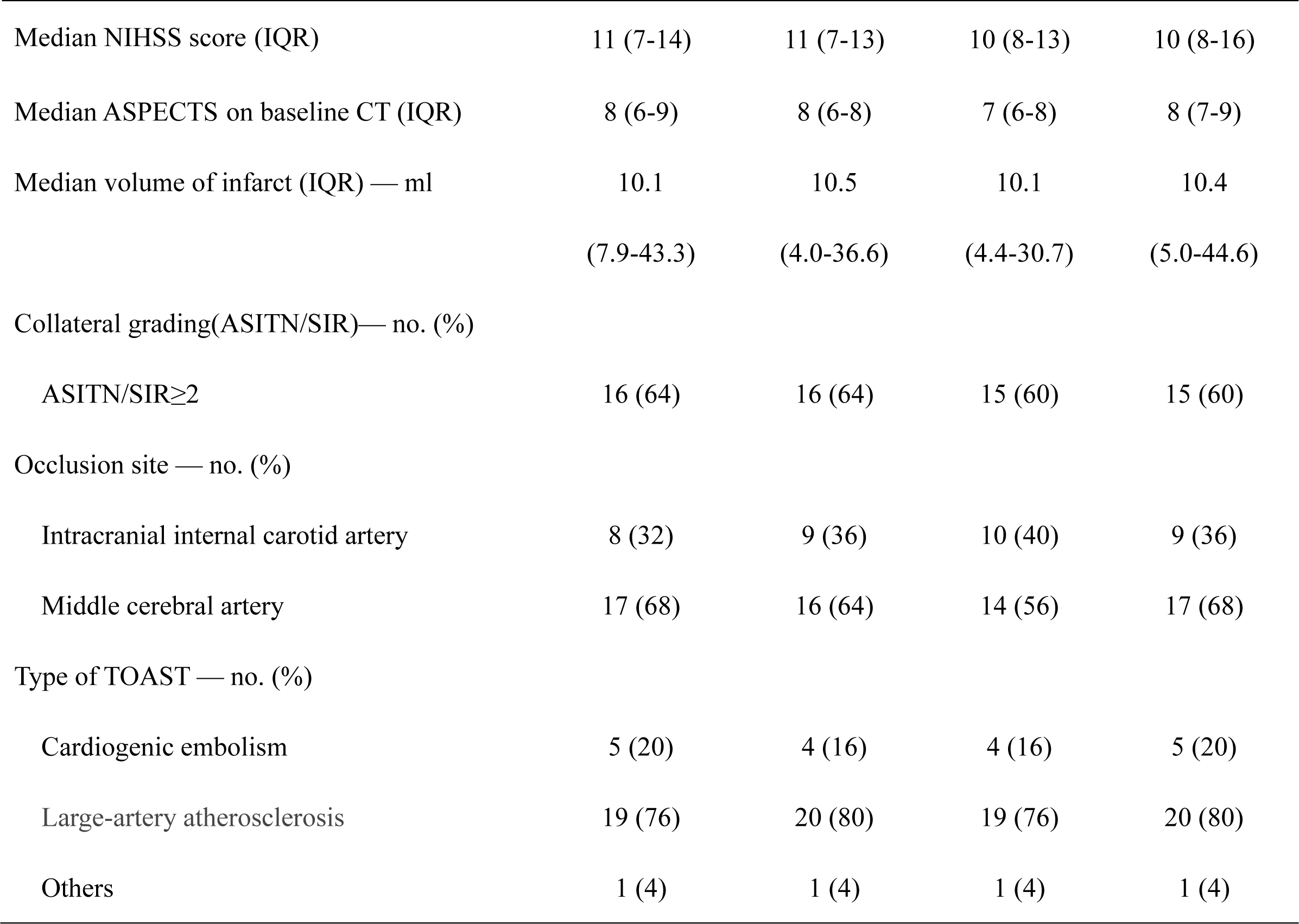
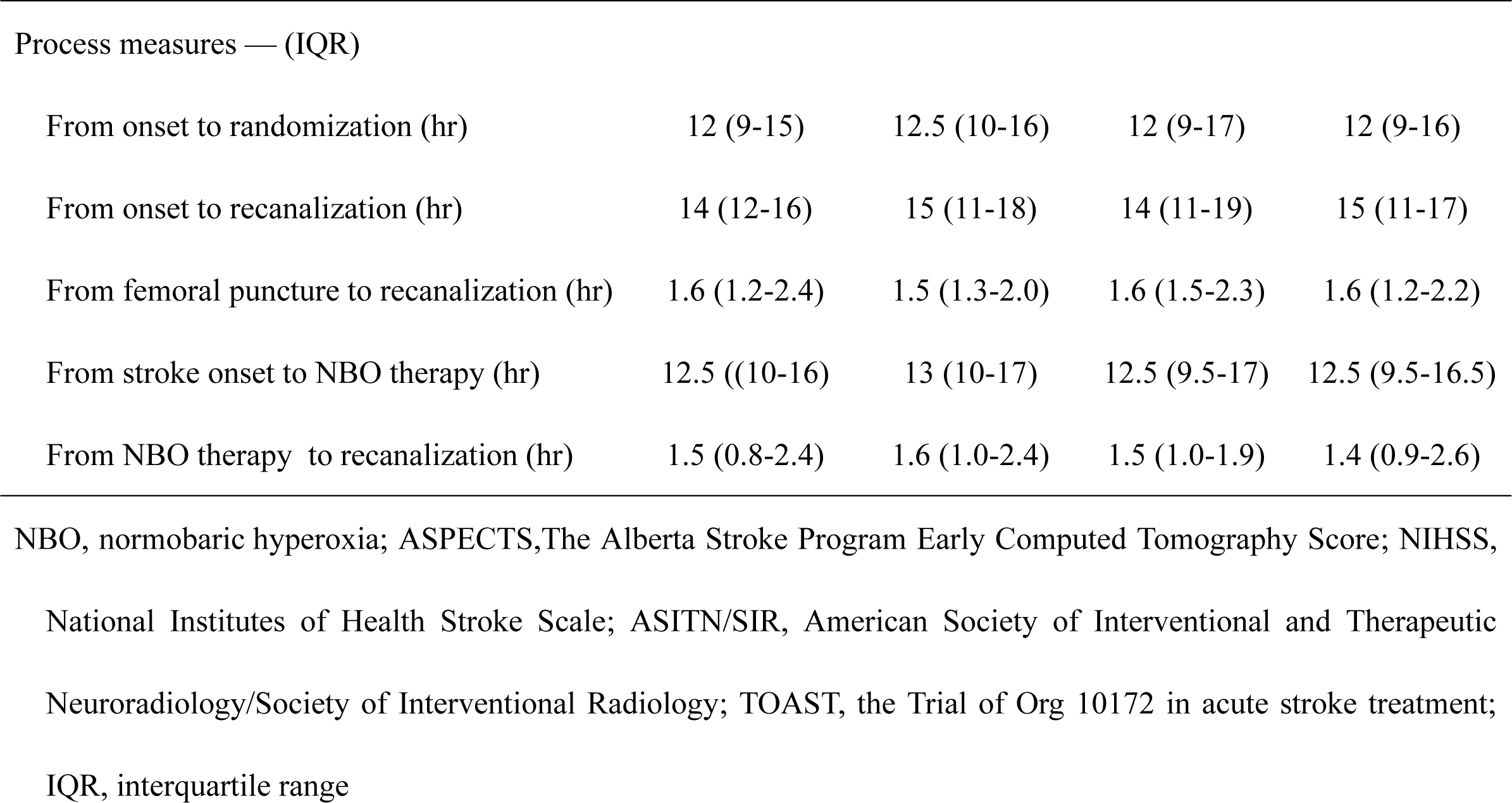
Characteristics of the Patients at Baseline.

| **Variable** | **Sham NBO**<br>**(n=25)** | **NBO-2h**<br>**(n=25)** | **NBO-4h**<br>**(n=25)** | **NBO-6h**<br>**(n=25)** |
| --- | --- | --- | --- | --- |
| Age — yr | 63.9 ± 9.6 | 61.6 ± 7.9 | 62.1 ± 8.3 | 62.0 ± 9.7 |
| Male sex — no. (%) | 13 (52) | 18 (72) | 16 (64) | 18 (72) |
| Atrial fibrillation — no. (%) | 5 (20) | 4 (16) | 5 (20) | 5 (20) |
| Diabetes mellitus — no. (%) | 6 (24) | 6 (24) | 6 (24) | 6 (24) |
| Hypertension — no. (%) | 16 (64) | 14 (56) | 14 (56) | 16 (64) |
| Previous ischemic stroke — no. (%) | 5 (20) | 6 (24) | 4 (16) | 4 (16) |
| PaO2 (mm Hg) at baseline— Mean (SD) | 91.0 ± 10.3 | 91.5 ± 14.3 | 90.7 ± 12.6 | 92.0 ± 10.5 |
| Median NIHSS score (IQR) | 11 (7-14) | 11 (7-13) | 10 (8-13) | 10 (8-16) |
| Median ASPECTS on baseline CT (IQR) | 8 (6-9) | 8 (6-8) | 7 (6-8) | 8 (7-9) |
| Median volume of infarct (IQR) — ml | 10.1<br>(7.9-43.3) | 10.5<br>(4.0-36.6) | 10.1<br>(4.4-30.7) | 10.4<br>(5.0-44.6) |
| Collateral grading(ASITN/SIR)— no. (%) |  |  |  |  |
| ASITN/SIR≥2 | 16 (64) | 16 (64) | 15 (60) | 15 (60) |
| Occlusion site — no. (%) |  |  |  |  |
| Intracranial internal carotid artery | 8 (32) | 9 (36) | 10 (40) | 9 (36) |
| Middle cerebral artery | 17 (68) | 16 (64) | 14 (56) | 17 (68) |
| Type of TOAST — no. (%) |  |  |  |  |
| Cardiogenic embolism | 5 (20) | 4 (16) | 4 (16) | 5 (20) |
| Large-artery atherosclerosis | 19 (76) | 20 (80) | 19 (76) | 20 (80) |
| Others | 1 (4) | 1 (4) | 1 (4) | 1 (4) |
| Process measures — (IQR) |  |  |  |  |
| From onset to randomization (hr) | 12 (9-15) | 12.5 (10-16) | 12 (9-17) | 12 (9-16) |
| From onset to recanalization (hr) | 14 (12-16) | 15 (11-18) | 14 (11-19) | 15 (11-17) |
| From femoral puncture to recanalization (hr) | 1.6 (1.2-2.4) | 1.5 (1.3-2.0) | 1.6 (1.5-2.3) | 1.6 (1.2-2.2) |
| From stroke onset to NBO therapy (hr) | 12.5 ((10-16) | 13 (10-17) | 12.5 (9.5-17) | 12.5 (9.5-16.5) |
| From NBO therapy to recanalization (hr) | 1.5 (0.8-2.4) | 1.6 (1.0-2.4) | 1.5 (1.0-1.9) | 1.4 (0.9-2.6) |

NBO, normobaric hyperoxia; ASPECTS, The Alberta Stroke Program Early Computed Tomography Score; NIHSS, National Institutes of Health Stroke Scale; ASITN/SIR, American Society of Interventional and Therapeutic Neuroradiology/Society of Interventional Radiology; TOAST, the Trial of Org 10172 in acute stroke treatment; IQR, interquartile range

### Safety outcomes

Two deaths (8%) occurred in the NBO-2h group, with one death (4%) occurring in each of the remaining groups. However, the difference in mortality rates was not significant. Two deaths (one in the Sham-NBO group and one in the NBO-2h group) were attributed to stroke-related causes (Table 2).

**Table 2.** Safety outcomes.

| Outcomes | Sham NBO<br>(n=25) | NBO-2h<br>(n=25) | NBO-4h<br>(n=25) | NBO-6h<br>(n=25) | P Value<sup>*</sup> |
| --- | --- | --- | --- | --- | --- |
| Death from any cause at 90 days— no. (%) | 1 (4) | 2 (8) | 1(4) | 1(4) | 0.935 |
| Stroke-related death at 90 days — no. (%) | 1 (4) | 1 (4) | 0 | 0 | 0.737 |
| Symptomatic intracranial hemorrhage (sICH) — no. (%) | 1 (4) | 2 (8) | 2 (8) | 1 (4) | 0.808 |
| Any intracerebral hemorrhage transformation — no. (%) | 5 (20) | 7 (28) | 7 (28) | 6 (24) | 0.874 |
| Any symptoms of discomfort after oxygen inhalation— no. (%) | 0 | 0 | 0 | 0 | NA |
| Early neurologic deterioration— no. (%) | 1(4) | 2 (8) | 1(4) | 1(4) | 0.935 |
| Serious adverse event (SAE)— no. (%) | 6 (24) | 5 (20) | 4 (16) | 6 (24) | 0.948 |
| Pneumonia— no. (%) | 4 (16) | 6 (24) | 4 (16) | 5 (20) | 0.918 |

\* The p value was calculated using the Fisher exact test.

The incidence of sICH was 4% in the Sham-NBO and NBO-6h groups (one case in each group) and 8 % in the NBO-2h and NBO-4h groups (two cases in each group). However, these differences were not significant (Table 2).

The incidence of any intracranial hemorrhage were as follows: 20% in the Sham-NBO group, 28% in the NBO-2h group, 28% in NBO-4h group, and 24% in the NBO-6h group. However, these differences were not significant. Notably, no cases of oxygen-related adverse symptoms were detected in any of the groups (Table 2).

END occurred in all the groups with two cases observed in the NBO-2-h group and one case observed in each of the remaining groups. Severe adverse events occurred in six cases in both the Sham-NBO (24%) and NBO-6h (24%) groups, with five cases (20%) reported in the NBO-2h group, and four (16%) in the NBO-4h group. Notably, no significant differences were observed among the four groups in terms of the incidence of pneumonia (Table 2).

### Efficacy outcomes

#### Primary efficacy outcomes

Infarct volume at 72 h, measured by MRI-DWI, was as follows: 39.4 ± 34.3 mL in the Sham-NBO group, 30.6 ± 30.1 mL in the NBO-2h groups, 19.7 ± 15.4 mL in the NBO-4h group, and 22.6 ± 22.4 mL in the NBO-6h group. The differences in infarct volume among the groups were significant (P=0.013, Figure 2, Table 3). Pairwise comparisons showed significant differences in the 72 h infarct volume of the NBO-4h and NBO-6h groups when compared to the Sham-NBO group (P=0.006 and P=0.005, respectively), with adjusted P-values of 0.011 and 0.027, respectively. However, no significant differences were observed between the NBO-4h and NBO-6h groups (P=0.702). A significant difference was observed in the 72 h infarct volume between the NBO-2h group and the Sham-NBO group before adjustment (P=0.045), but this difference was not observed significantly after adjustment (P=0.271).

**Figure 2.**
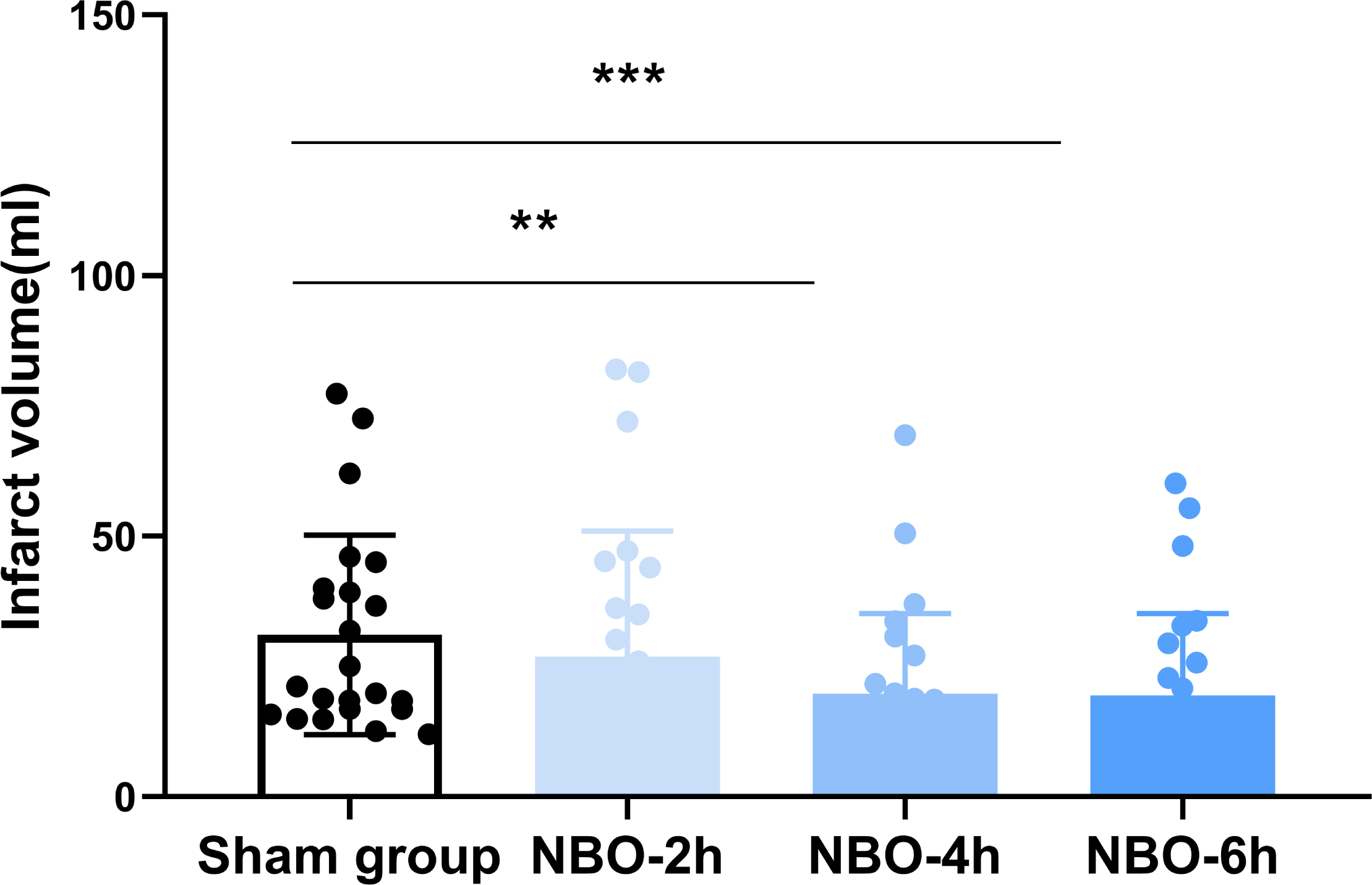
Comparison of cerebral infarction volume. The infarction volumes were 39.4 ± 34.3 ml for the low-flow oxygen group, 30.6 ± 30.1 ml for the 2-hour NBO group, 19.7 ± 15.4 ml for the 4-hour NBO group, and 22.6 ± 22.4 ml for the 6-hour NBO group. A significant difference was observed among the four groups (P=0.013). Pairwise comparisons revealed that both the 4-hour NBO group and the 6-hour NBO group showed significant statistical differences compared to the Sham-NBO group (corrected P values were 0.011 and 0.027, respectively).

**Table 3.**
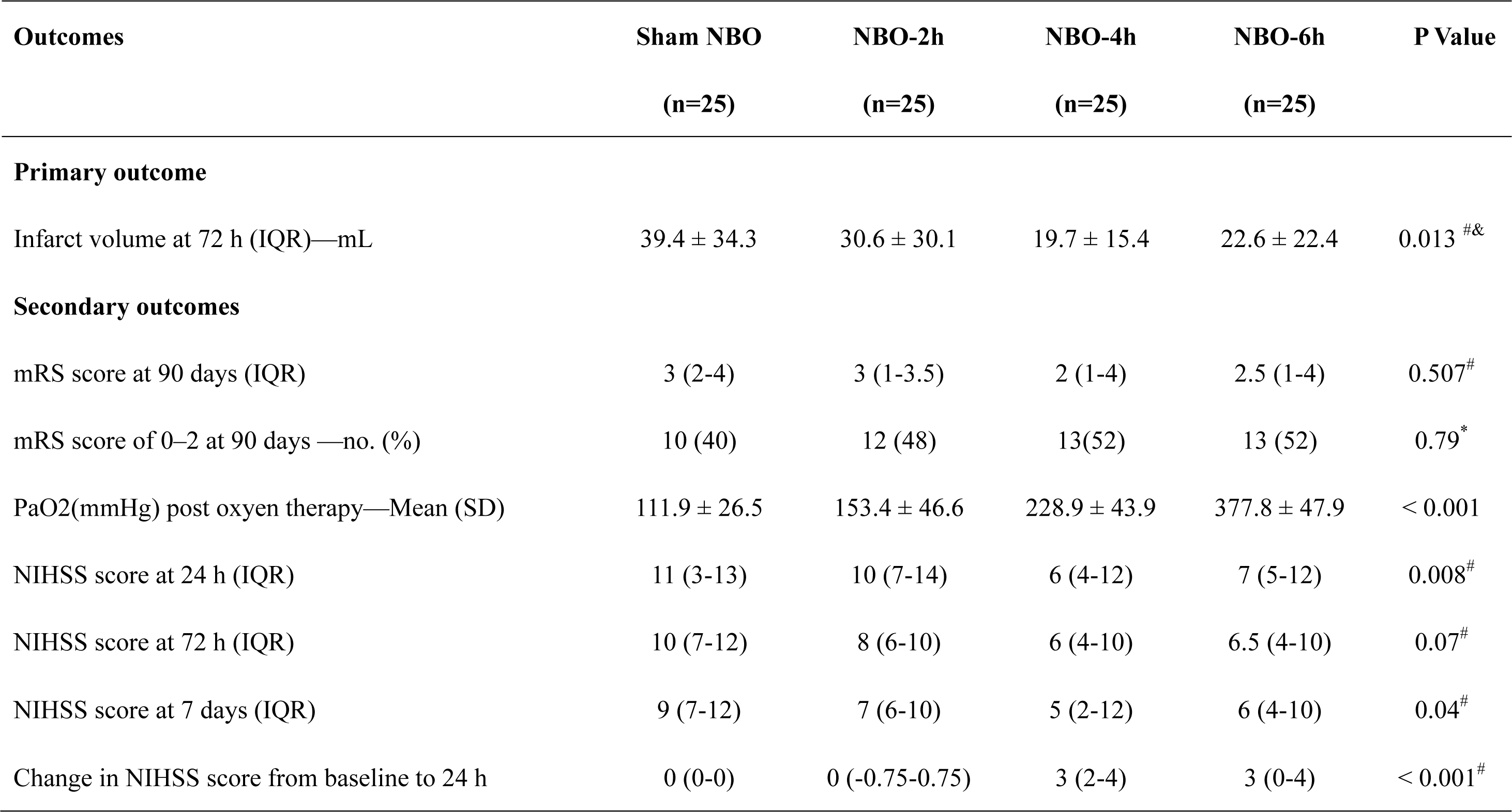
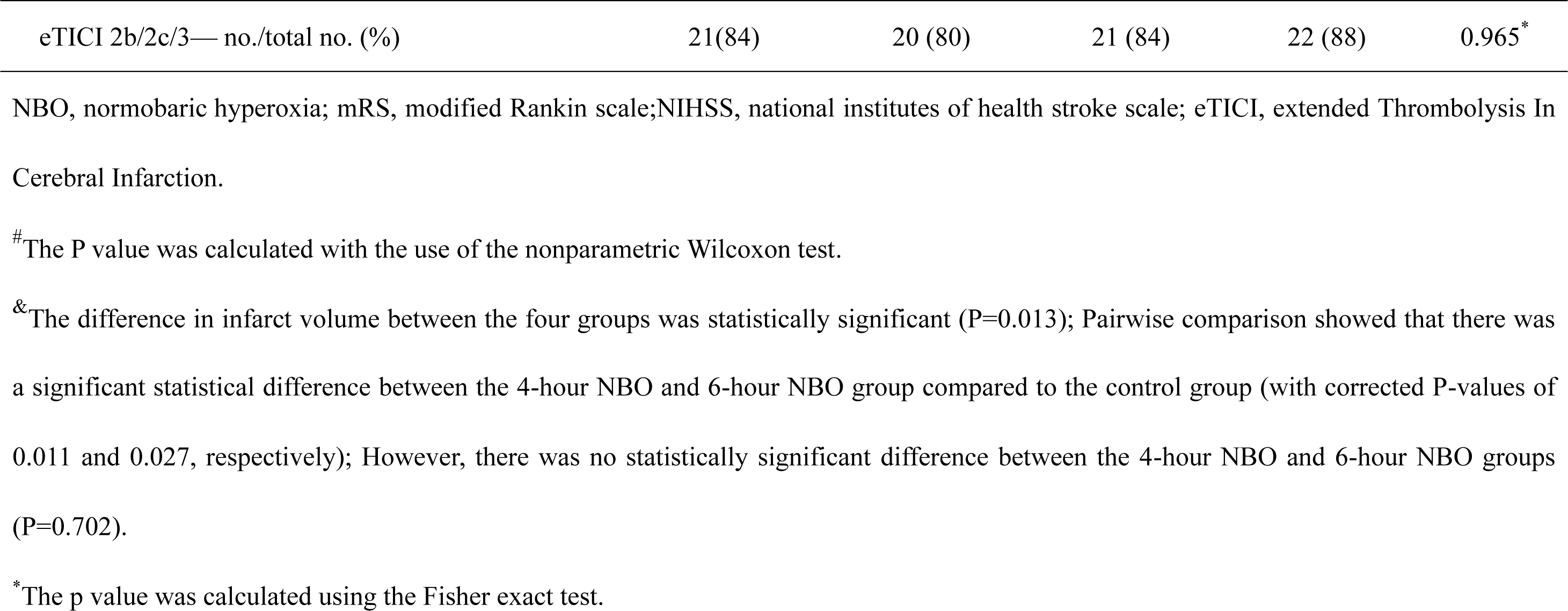
Efficacy Outcomes.

#### Secondary Efficacy Outcomes

The median (IQR) 90-d mRS score was 3 (2-4) in the Sham-NBO group, 3 (1-3.5) in the NBO-2h group, 3 (1-3.5) in the NBO-4h group, and 3 (1-3.5) in the NBO-6h group. These differences were not significant (Table 3).

The rate of favourable outcomes at 90 d was the highest in the NBO-4h group (52%) and the NBO-6h group (52%), with the lowest rate in the Sham-NBO group (40%). However, these differences were not significant(Table 3, Figure 3).

**Figure 3.**
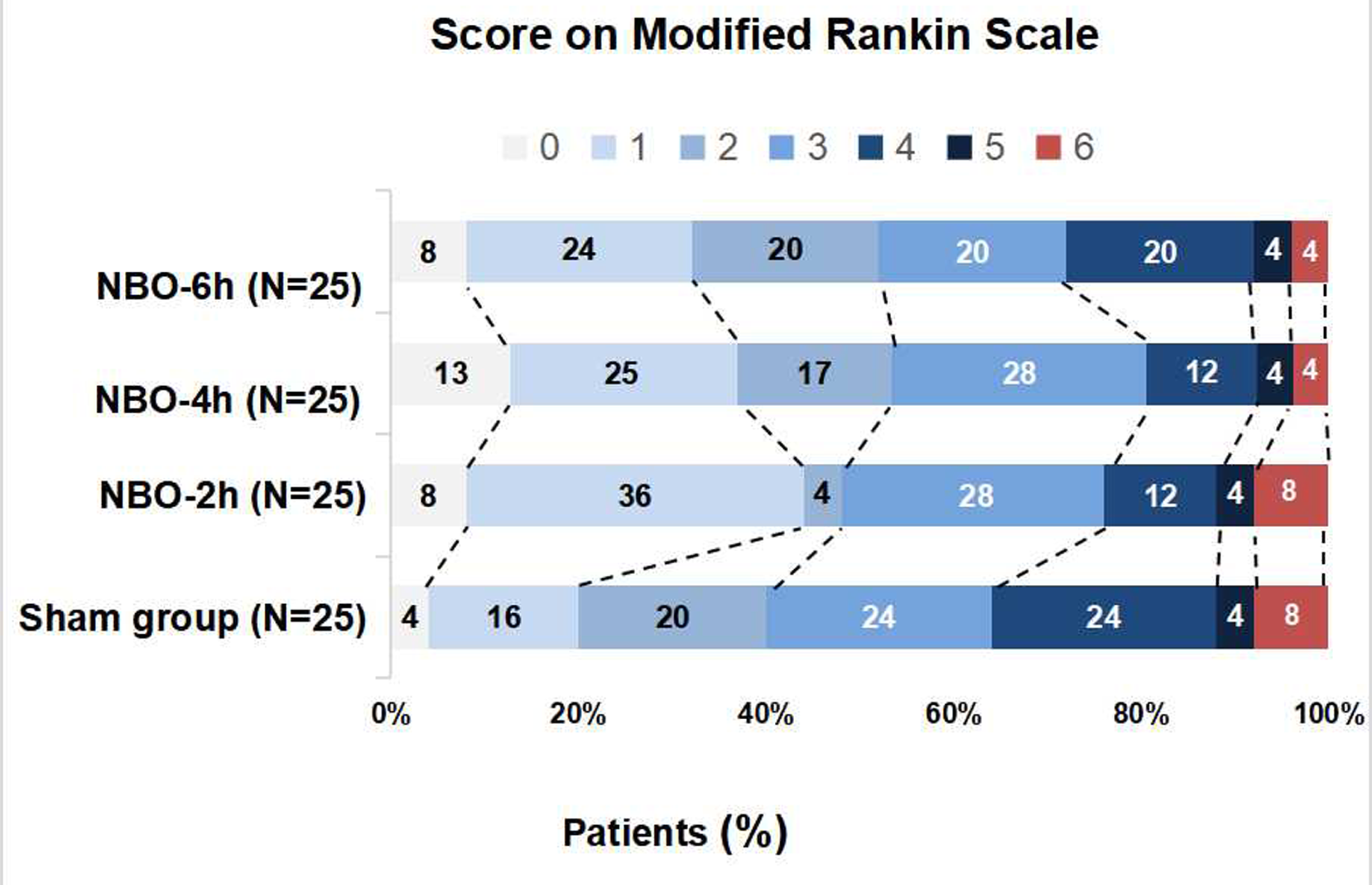
Distribution of the modified Rankin scale scores at 90 days. The rate of favourable outcomes (mRS0-2) at 90 days was the highest in the NBO-4h group (52%) and the NBO-6h group (52%), with the lowest rate in the Sham-NBO group (40%). However, these differences were not significant.

An increase in the PaO_2_ was observed with the increase in duration of oxygen therapy as follows: PaO2 was 111.9 ± 26.5 mmHg in the Sham-NBO group, 153.4 ± 46.6 mmHg in the NBO-2h group, 228.9 ± 43.9 mmHg in the NBO-4h group, and 377.8 ± 47 mmHg in the NBO-6h group. A significant difference was observed in oxygen tension levels among the four groups (P<0.001, Table 3).

A significant difference was observed in the median 24 h NIHSS scores among the four groups (p=0.008, Table 3). In pairwise comparisons, significant differences were observed in the median 24 h NIHSS scores in comparison to both the NBO-4h [6 (IQR 4-12)] and the NBO-6h [7 (IQR 5-12)] groups with the Sham-NBO [11 (IQR 3-13)] (P=0.007 and P=0.010, respectively) and the NBO-2h groups [10 (IQR 7-14)] (P=0.02 and P=0.029, respectively). However, no significant differences were observed between the NBO-2h group and the Sham-NBO group or between the NBO-4h group and the NBO-6h group.

The improvement in the median 72 h NIHSS score in the NBO-4h [6 (IQR 4-10)] and NBO-6h [6.5 (IQR 4-10)] groups was higher than that in the Sham-NBO [10 (IQR 7-12)] and NBO-2h [8 (IQR 6-10)] groups; however, these differences were not significant (P>0.05,Table 3).

Significant differences in the median 7-day NIHSS score were observed among the four groups (P=0.04, Table 3). Pairwise comparisons indicated that both the NBO-4h [5 (IQR 2-12)] and NBO-6h [6 (IQR 4-10)] groups exhibited significant differences compared to the Sham-NBO group [9 (IQR 7-12)] (P=0.018 and P=0.027, respectively). However, no significant differences were found between the NBO-2h group and the Sham-NBO group (P>0.05). Furthermore, no significant differences were found between the NBO-4h group and NBO-6h group (P>0.05).

A significant difference was observed among the four groups in the changes in median NIHSS scores from baseline to 24 h (P<0.001, Table 3). In pairwise comparisons, both the NBO-4h group [3 (IQR 2-4)] and the NBO-6h group [3 (IQR 0-4)] showed significant differences compared to the Sham group [0 (IQR 0-0)] (P<0.01) and the NBO-2h group [0 (IQR −0.75-0.75)] (P<0.01). However, no differences were found between the NBO-2h group and the Sham-NBO group or between the NBO-4h group and the NBO-6h group.

Regarding the rate of successful vascular recanalization (eTICI 2b/2c/3), all four groups had a recanalization rate of over 80% with no significant differences among the four groups (Table3).

## DISCUSSION

In the present study, a specialized storage oxygen mask was used, which allows delivery of higher concentrations of oxygen compared to regular masks. Various time gradients (2h, 4h, and 6h) were established, and a randomized controlled trial was conducted to investigate the most effective treatment duration of NBO. This study revealed a significant reduction in infarct volume in both the NBO-4h and NBO-6h groups compared to that in the Sham-NBO group. Importantly, no significant difference in infarct volume between the NBO-4h group and NBO-6h group. Additionally, no significant differences were found at baseline among the groups, which enhances the comparability of the groups and supports the robustness of the results.

Previous clinical studies on the investigation of NBO therapy applied different treatment regimens.^16, 24, 25^ Singhal et al. applied NBO therapy consistently for 8 h at a rate of 45 L/min by a simple face mask and reported a beneficial neuroprotective effect for 1 week although treatment was found to be ineffective at 90 days.^16^ Padma et al. found NBO therapy for 12 h at 10 L/min by a simple face mask to be ineffective ^25^; Mazdeh et al. found that NBO therapy via a Venturi mask can improve long time outcomes in the patients who have either ischemic or haemorrhagic stroke with an oxygen saturation set at 50% and continuous oxygen inhalation for 12 hours.^26^ Similarly, Cheng et al. administered NBO via a Venturi mask (FiO2, 50%; flow, 15 L/min) for 6 h after vessel recanalization concluding that high-flow NBO therapy after endovascular recanalization was safe and effective in improving functional outcomes, decreasing mortality, and reducing infarct volumes in patients with anterior circulation stroke within 6 h from stroke onset.^18^ Furthermore, the PROOF study, an ongoing study by Poli et al., investigated the application of NBO before vessel recanalization, with results pending disclosure.^17^ Our team conducted the OPENS-1 study, in which NBO therapy was administered in multiple sequential stages—namely, before, during, and after vessel recanalization—using a continuous storage oxygen mask at 10 L/min for 4 h, demonstrating both the effectiveness and safety of treatment.^14^ Notably, these studies vary in terms of study populations, ethnicities, and the types of oxygen masks employed so comparisons of results need to be approached with caution.

Henninger et al. established a model of permanent focal cerebral ischemia to investigate the treatment effect of NBO at different durations of 3 h and 6 h, which showed that NBO therapy for 6 resulted in a significantly better neuroprotection effect, with a 44% reduction in cerebral infarct volume compared to the normoxia group.^27^ This effect was reduced by 10% with 3 h of NBO therapy indicating that increased durations of NBO therapy offer improved neuroprotective effects.

Our study indicated that both 4 h and 6 h of NBO therapy were more effective in reducing infarct volume and improving neurological function than either low flow oxygen therapy or 2 h of NBO therapy. However, no significant differences were observed in effectiveness between the 4-h and 6-h NBO groups. Regarding safety outcomes, no significant differences were observed among the 2-, 4-, and 6-h NBO treatment groups. Considering both effectiveness and safety, these results suggest that NBO therapy for 4-6 h is feasible and provides benefits in terms of reducing infarct volume and improving neurological outcomes. However, from an economic and patient tolerance perspective, a shorter duration of oxygen therapy may be more favourable. Therefore, to maximise the benefits of NBO therapy and patient tolerance and compliance, NBO therapy for 4 h may offer a balanced approach in terms of patient acceptance and cost-effectiveness.

This study has several strengths. First, we investigated the effectiveness and safety of varying durations of NBO treatment for AIS through a randomised controlled design. This approach enhances the objectivity of the results and generates robust evidence. Second, the study aimed to determine the optimal duration of NBO treatment in the context of recanalizing the occluded blood vessel, which distinguishes it from previous research ^16, 24, 25^ that predominantly focused on patients with stroke without vessel recanalization. Furthermore, while earlier studies ^27, 28^ often relied on animal experiments to explore NBO temporal gradients, this study examined NBO temporal gradients in patients with stroke with results that are more directly applicable to clinical practice.

### Limitations

This study is subject to several noteworthy limitations: First, the study was a single centre study with potential inherent statistical biases. Second, the study investigated the delivery of NBO within the 2 to 6-h timeframe. Future inquiries should examine extending the delivery of NBO over a more prolonged duration, such as 8-10 h. However, caution when extending the duration of NBO therapy is imperative, as this may amplify the risk of NBO-associated adverse effects, such as increasing vulnerability to oxidative stress. Finally, the primary endpoint of this study was the assessment of the 72-h infarct volume. Future investigations should examine for more reliable endpoints, including pre- and post-NBO therapy changes in the volume of the penumbra and functional outcomes.

In summary, the efficacy of NBO varied according to the duration of treatment. NBO treatment for 4-6 h following successful revascularization was more effective than treatment for 2 hours or no oxygen treatment. Our study findings offer valuable insights into the clinical use of NBO and its integration into the routine management of patients with stroke.

## Data Availability

Any data not published within the article will be shared on request from any qualified investigator.

## Acknowledgments

We would like to thank Editage (www.editage.cn) for English language editing.

## Sources of funding

This study was sponsored by the Talent Construction Fund of Beijing Institute of Brain Disorders (PXM2020_014226_000004), the National Natural Science Foundation of China (82101389), and Tianjin Key Research and Development Program in Science and Technology (19YFZCSY00260).

## Disclosures

None.

## Non-standard Abbreviations and Acronyms

AIS: acute ischemic stroke
NBO: normobaric hyperoxia
EVT: endovascular treatment
mRS: modified Rankin Scale
FiO2: inspiratory oxygen fraction
ADC: apparent diffusion coefficient
PaO2: partial pressure of oxygen
eTICI: extended Thrombolysis in Cerebral Infarction scale
NIHSS: National Institutes of Health Stroke Scale
sICH: Symptomatic intracerebral hemorrhage
END: Early neurological deterioration

## Notes

### Competing Interest Statement

The authors have declared no competing interest.

### Clinical Trial

URL: https://www.clinicaltrials.gov, Unique identifier: NCT05404373

### Author Declarations

The Ethics Committee of Huanhu Hospital, Tianjin Medical University (Approval No. 2022-076)

